# A Climate-Driven Mechanistic Transmission Model to Characterize Dengue Epidemiology in Dhaka, Bangladesh

**DOI:** 10.1101/2025.02.19.25322534

**Authors:** Kishor K Paul, David G Regan, Donna Green, Ian Macadam, Richard T Gray

## Abstract

Dengue is a major public health concern in tropical and sub-tropical areas of the world. Dengue virus is transmitted to humans primarily by female *Aedes aegypti* mosquitoes and its epidemiology is heavily influenced by the climate. We developed a mechanistic compartmental model to characterize how climate and the evolution of population-level immunity influence changes in the epidemiology of dengue. The model explicitly incorporates aquatic and adult stages of the mosquito vector and cross-immunity (to another serotype) to better represent the transmission of dengue compared to previous models. It incorporates parameters related to mosquito development, behaviour, and mortality that are dependent on temperature, rainfall, and humidity. We calibrated the model to available dengue seroprevalence estimates for Dhaka, Bangladesh as it is a well-observed densely populated endemic area for dengue and will be strongly impacted by climate change. We then simulated dengue epidemiology for the period of 1995-2014 and compared the simulation output with available epidemiologic data. The model produced repeating annual outbreaks of dengue with strong seasonality, as observed in the dengue case reports for Dhaka. The median number of annual dengue infections produced by the model was 1.6 million cases (IQR: 0.6-2.3 million) and the median of the annual daily peak size of dengue infections was 24,121 (IQR: 7,807-39,308) across the simulations. These figures while exceeding reported cases in Dhaka, which do not include asymptomatic infections or cases not seeking healthcare, align with previous estimates. The modelled seroprevalence of 62% (IQR: 58– 75%) aligned with survey results from Dhaka. In most model simulations, dengue infections occurred during August to December with the median peak occurring in October. The model outputs demonstrated a high degree of conformity to the reported data in terms of seasonality, and seroprevalence, indicating that the model provides a good representation of overall dengue epidemiology in Dhaka. By accounting for climate variables, host immunity, and their interplay, our model is well-suited for evaluating the long-term impact of climate change on dengue transmission in Dhaka and other settings where climate and epidemiological data are available.

## Introduction

Dengue, a mosquito-borne virus, causes about 390 million new infections each year globally, of which 96 million infections present as a clinically apparent disease of variable severity [1]. Most dengue virus infections are asymptomatic, but symptoms from mild fever to haemorrhagic shock and even death can occur if not managed properly. There are four genetically related but distinct dengue serotypes known as DENV-1, -2, -3, and -4 which affect dengue epidemiology [2]. The dengue virus is transmitted to humans by mosquitoes of the genus *Aedes*, most commonly *Ae. aegypti*. The lifecycle of the dengue mosquito vector is influenced by climate variables, and consequently so are dengue virus dynamics [3]. This sensitivity to climate, and the emergence of dengue as a public health issue, means that there is a need to assess how dengue epidemiology could be affected by climate change.

Both statistical and mathematical modelling approaches have been used to estimate dengue outbreak risk in the short-term (a few months to a few years) for given temperature, rainfall, and humidity data [4-6]. Recent studies employing statistical modelling and encompassing diverse climatic conditions have incorporated global dengue occurrence and mosquito vector data, to model dengue spread and future mosquito vector distribution and population at risk [7, 8]. However, these non-mechanistic models do not account for underlying causal factors influencing dengue transmission and observed epidemiological patterns, such as population-level immunity. This limits our confidence in their ability to assess the impact of future climate change on dengue.

Mechanistic models that do attempt to represent relevant causal relationships also have limitations. In particular, they require specific location data for calibration and validation which may not be readily available [9]; some models focus solely on the mosquito population dynamics, neglecting the transmission of dengue to human hosts [10]; and some models overlook the co-circulation of multiple dengue serotypes and resulting cross-immunity that shape long-term dengue epidemiology [11]. Additionally, mechanistic models used to assess the impact of climate change on the risk of dengue have generally focused on the effects of temperature, ignoring other climate variables. For example, vectorial capacity (VC) of *Ae. aegypti* mosquitoes is used as an indicator in the annual *Lancet Countdown* impact of climate change on health assessment but only uses temperature as an input [12-14]. However, changes in other climate variables such as rainfall and humidity are potentially important drivers of dengue risk [10, 13]. We identified only one published study that projected dengue epidemics up to the year 2070 and considered co-circulation of multiple dengue serotypes and the impact of changing temperature and rainfall [15]. To overcome some of these limitations, we developed a mechanistic model that considers both climate factors and host immunity. This model enables us to make long-term projections of dengue epidemiology in a changing climate.

Bangladesh and, in particular, its major cities, have high levels of dengue seroprevalence for multiple dengue serotypes [16]. This means immunity and cross-immunity likely play a crucial role in supressing dengue transmission. Additionally, climate models generally agree on a future increase in temperature and precipitation in Bangladesh, though there remains some uncertainty regarding the magnitude of these changes [17]. In this study, we applied our mechanistic model to Dhaka, the capital of Bangladesh. We selected Dhaka because of the availability of monthly records of dengue case numbers for a relatively long period, 2000-2014, and population-level seroprevalence estimates for 2014. We demonstrate that the model performs well at simulating recent dengue outbreak characteristics, suggesting it is suitable for use in assessments of the long-term impact of climate change on dengue epidemiology.

## Methods

### Epidemiological and demographic data

During the period in which this research was conducted, we had access to monthly reports of dengue cases and serotype distribution for Dhaka for the period 2000-2014 from the Institute of Epidemiology, Disease Control and Research, Ministry of Health and Family Welfare, Government of Bangladesh [18]. The number of annual reported dengue cases in Dhaka ranged from 6,232 in 2002 to 375 in 2014 (Figure 1A). This reflects only an estimated 2.8% of all annual dengue cases in the city, primarily the severe cases that required hospitalization [19]. Despite this, the data provided insight into the relative scale and highly seasonal nature of dengue outbreaks from one year to the next, with cases starting to rise each July, peaking during September, and then dropping to near zero by the end of December [18].

**Figure 1:**
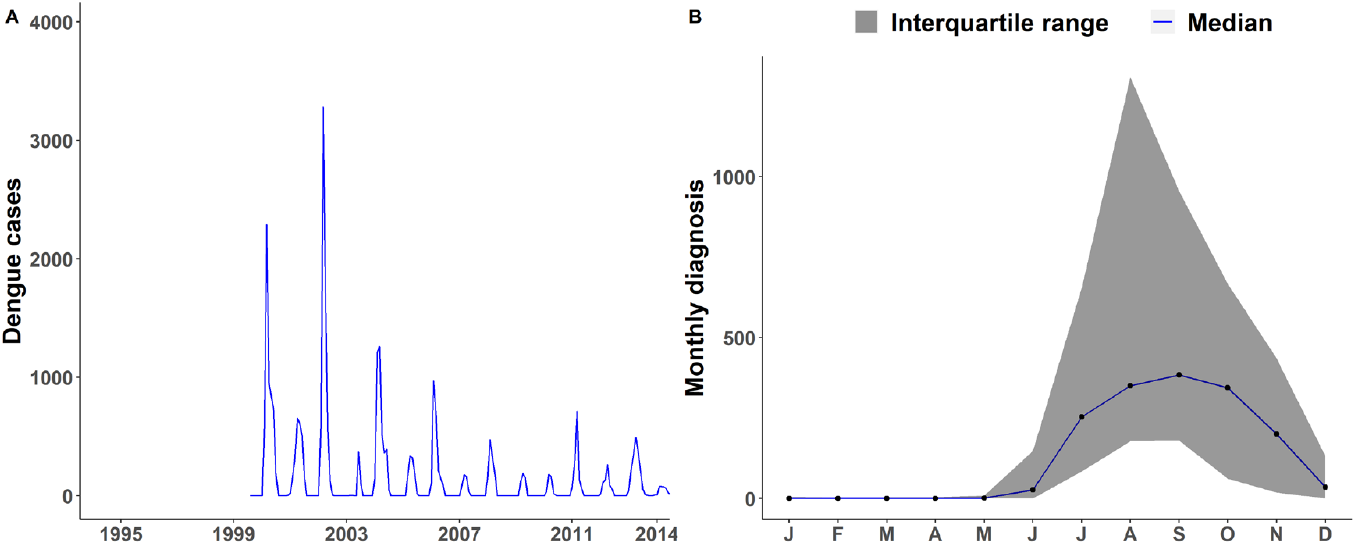
Reported dengue cases in Dhaka, Bangladesh (2000 – 2014) (A) Reported monthly number of cases. (B) Monthly distribution of reported cases. Data sourced from the Institute of Epidemiology, Disease Control and Research, Ministry of Health and Family Welfare, Government of Bangladesh [18].

A previous study that analysed blood samples that were collected in 1996-1997 suggested that dengue was circulating prior to the outbreak in 2000 and estimated a seroprevalence of 0.5% within the population during that period [20]. Based on this epidemiological data, we introduced two dengue serotypes sequentially in our model, the first serotype in 1995, and the second in 2000. Two more recent studies provided seroprevalence estimates for Dhaka with one estimating a seroprevalence of 80% in 2012 and the other estimating a seroprevalence range of 26-85% in 2014 [21, 22].

To account for change in the human population of Dhaka from 1995 to 2014, we used data on human birth rates, death rates, and population movement [23, 24]. We obtained the crude birth and death rates for Bangladesh from the United Nation Population Division Data Portal and applied these rates as best available estimates for Dhaka [25].

### Observed climate data

Dhaka has one meteorological observing station at Lat 23°47’N, Long 90°23’E for which temperature, relative humidity, and rainfall data are available from the Bangladesh Meteorological Department, [26]. We obtained daily records of temperature (mean, minimum, and maximum), relative humidity, and total rainfall logged at the observing station for the period 1995-2014. The choice of this timeframe for climate data was guided by the availability of comprehensive monthly dengue case reports up to 2014, and indications of emergence of dengue in Dhaka around 1995 [20].

### Dengue transmission model

We developed a new compartmental model of dengue transmission, adapting previous approaches to explicitly incorporate the impact of climate variables on aquatic and adult stages of the mosquito vector and cross-immunity in the human population, with a view to better represent dengue dynamics at the population level over the long-term.

For the human population, we expanded a previously developed two strain-dengue model to incorporate cross-immunity to different dengue serotypes [15]. Although there are four dengue serotypes, for simplicity, we chose to incorporate just two serotypes of dengue in our model to represent circulation of any two of the four serotypes. Previous research has demonstrated that someone who survives two consecutive infections with different dengue serotypes acquires immunity against all four serotypes, suggesting that modelling more than two serotypes is unnecessary [27]. After being infected with any of the serotypes, an individual initially has cross-protective but waning immunity to the other serotype, but the immunity persists for the infecting serotype.[15] In the model, humans are born into the modelled population (Figure 2) susceptible (S_h_) to dengue infection after which they may be exposed (E_h1_ and E_h2_; infected but not yet infectious) to either dengue virus serotype by infectious bites from mosquitoes in the infected stages (I_v1_ and I_v2_) at a rate β_vh_, and then move to the infectious (I_h1_ and I_h2_) states and later to the cross-protection states (1°CI_h1_ and 1°CI_h2_). Individuals who have recovered from primary infection (R_h1_ and R_h2_) become susceptible to infection with the second serotype and may be exposed (E_h12_ and E_h21_) to the second dengue serotype by bites from infectious mosquitoes at a rate β_vh_. As β_vh_ represents the rate of transmission between an infected vector and a susceptible host, we assumed this rate to be the same for both primary and secondary infections. Individuals with exposure to a second dengue serotype then move into the secondary infectious state (I_h12_ and I_h21_). In this state, they are more infectious to mosquitoes and experience higher disease-induced mortality because of a process known as the antibody-dependent enhancement (ADE), where antibodies from the primary infection bind to second dengue serotype [28]. Seropositive and seronegative individuals can migrate into and out of the population. Seronegative immigrants enter the susceptible state, and we assumed that seropositive immigrants are not currently infected and are proportionately distributed among the recovered compartments at the time of entry. Parameters relevant to dengue virus transmission in the human population are listed in Table 1.

**Table 1:** Summary of climate independent model parameters related to dengue transmission in human hosts and mosquito vectors. To obtain sample values for parameters following a gamma or exponential distribution, we sampled from the respective distribution using the average and the truncated range (sampling range). Setting-specific parameters such as population demographics are described in Table 3.

| Symbol | Explanation | Median Value | Sampling Range, Distribution | Source |
| --- | --- | --- | --- | --- |
| $1/\alpha_1$ | Latent period for dengue virus type 1 in humans | 5.9 days | 3-10 days, Gamma | [42, 43] |
| $1/\alpha_2$ | Latent period for dengue virus type 2 in humans | 5.9 days | 3-10 days, Gamma | [42] |
| $1/\gamma_1$ | Infectious period for dengue virus type 1 in humans | 5 days | 2-14 days, Gamma | [11, 44, 45] |
| $1/\gamma_2$ | Infectious period for dengue virus type 2 in humans | 5 days | 2-14 days, Gamma | [11, 44] |
| $1/\sigma_1$ | Number of days a person remains cross immune to reinfection by dengue type 2 | 730 days | 365-1095 days, Exponential | [42, 46] |
| $1/\sigma_2$ | Number of days a person remains cross immune to reinfection by dengue type 1 | 730 days | 365-1095 days, Exponential | [46] |
| $\mu_{ha}$ | Disease induced human mortality rate | 0.005 per day | Fixed | [47] |
| ADE | Multiplicative effect of antibody dependent enhancement on transmission probability | 1.5 | Fixed | [28] |
| $v$ | Vertical infection rate of *Ae. aegypti* mosquitoes | 0.004 | Fixed | [48] |
| $\beta_{im}$ | Mosquito-to-human transmission multiplication factor | 0.3 | Fixed | Calibrated |
| $\beta_{mi}$ | Human-to-mosquito transmission multiplication factor | 0.3 | Fixed | Calibrated |
| $f$ | Fraction of eggs that can become female mosquitoes | 0.5 | Fixed | Assumption |
| $N_v/N_h$ | Initial female mosquito to human population ratio | 2 | Fixed | Assumption based on [15] |

**Figure 2:**
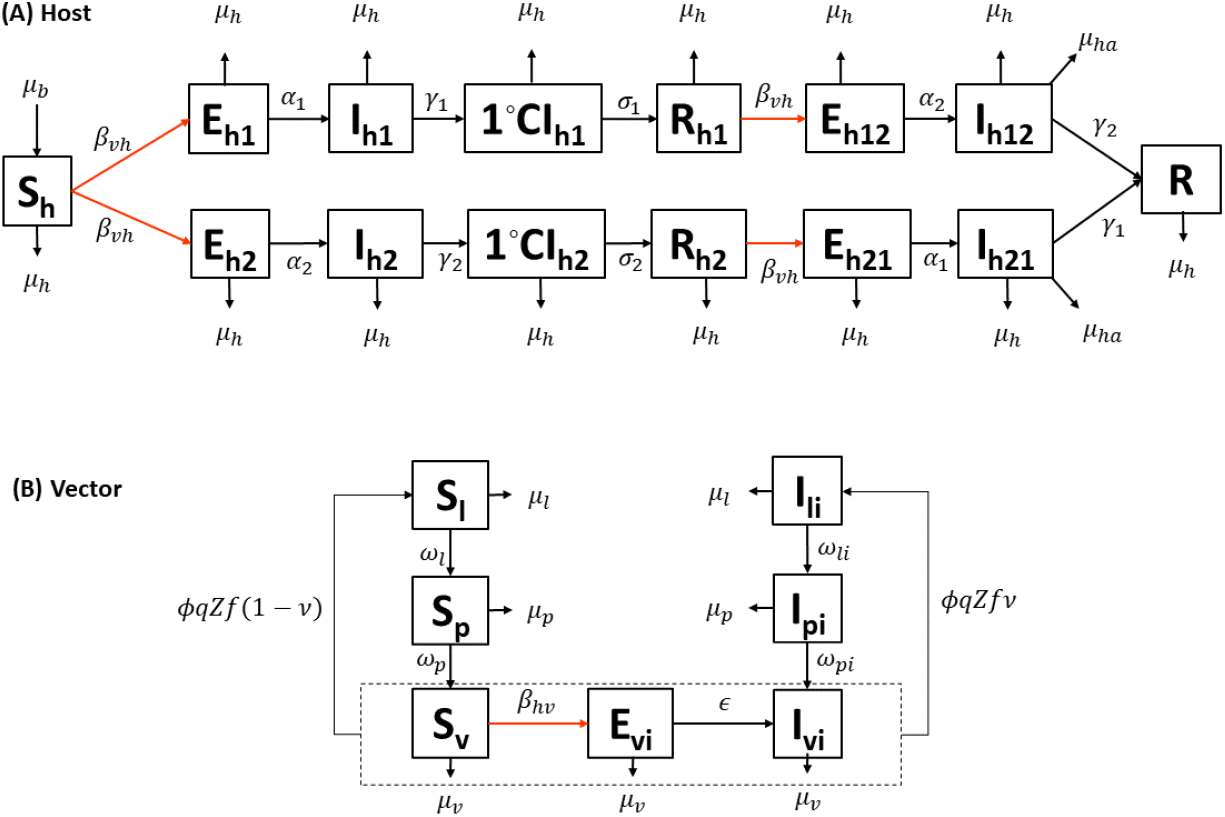
Epidemiological model framework. Two-serotype (i = 1, 2) dengue transmission model involving a human host population and a mosquito vector population; the human population is distributed between the susceptible (S_h_), exposed to first or second serotype (E_h1_ or E_h2_), infectious with first or second serotype (I_h1_ or I_h2_), primary cross-immune to one serotype (1°CI_h1_ or 1°CI_h2_), recovered from one serotype (R_h1_ or R_h2_), exposed to the heterogenous second serotype (E_h12_ or E_h21_), infectious with second serotype (I_h12_ or I_h21_), and recovered from both serotypes (R) states (compartments); arrows indicate the direction individuals can move out of compartments and red arrows indicate virus transmission events; transitions between compartments are labelled with the corresponding rate parameters; immigration into and emigration out of the modelled human population are not shown but are included in the model (see Equations 1-8 in the main text); the mosquito larva population is distributed between the susceptible (S_l_), and infectious (I_l_) states; the pupa populations is also distributed between the susceptible (S_p_), and infectious (I_p_) states; the adult mosquito population is distributed between the susceptible (S_v_), exposed (E_v_), and infectious (I_v_) states.

As mosquitoes have a relatively short lifespan in comparison to their human hosts, we assumed that mosquitoes can be infected with only one of the two dengue serotypes during their lifetime. Female mosquitoes become infected by feeding on human hosts who are either experiencing a primary or secondary dengue infection. We represent the mosquito lifecycle through the larva, pupa, and adult stages because these are the stages most affected by climate [10]. Larvae hatched from mosquito eggs can be either susceptible (S_l_) or infected by trans-ovarian transmission (I_l1_ and I_l2_). Susceptible and infected larvae can then progress to the pupa stage (S_p_, I_p1_ and I_p2_). The adult stage is divided into susceptible (S_v_), exposed (E_v1_ and E_v2_), and infectious (I_v1_ and I_v2_) states. In addition to vertical infection, mosquito vectors are exposed to dengue virus by biting infected human hosts in the infected stages (I_h1_, I_h2_, I_h12_, and I_h21_) with primary or secondary infection at a rate β_hv_. The parameters β_vh_ and β_hv_ capture the overall effects of transmission between mosquitoes and humans and could vary in different settings depending on variables not explicitly captured in the model.

The model is formulated as a system of 15 ordinary differential equations (Eq. 1-15 below). In these equations, subscript i = 1 or 2, denotes dengue virus serotype, and subscript ij denotes infection by serotype i preceding an infection by serotype j. The equations are coded as difference equations in R (version 4.1.1). All model code is available under an open access license at the following URL (https://github.com/kkpaul-ide/Dengue_Climate_Model) [29].

### Human

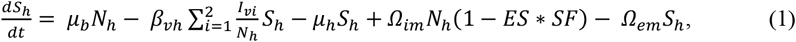

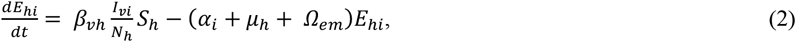

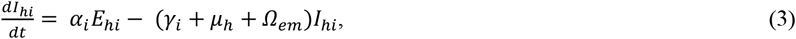

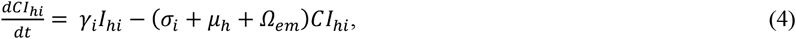

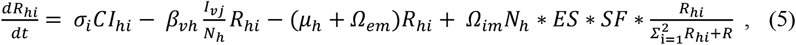

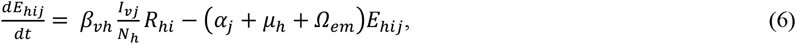

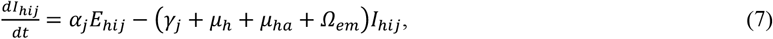

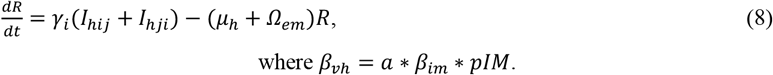

### Vector

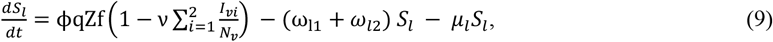

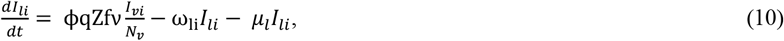

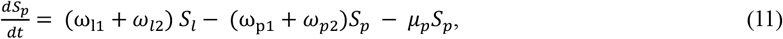

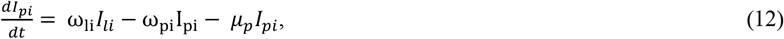

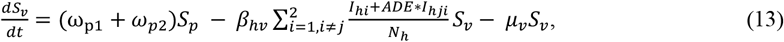

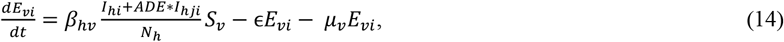

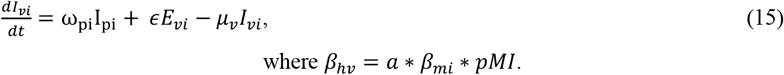

### Model input parameters

All parameters related to dengue infection in human hosts, the vertical infection rate, the fraction of eggs that can become female mosquitoes, the initial female mosquito to human population ratio, and the multiplication factors for dengue transmission (human-to-mosquito (β_mi_) and mosquito-to-human (β_im_), used for calibration) are assumed to be climate independent (Table 1). We assumed no change in human host behaviour due to the change in climate.

Based on available empirical studies, the following mosquito parameters are assumed to depend on temperature only: the biting rate (a), the per-bite probability of infection from human to mosquito (pIM), the oviposition rate(the number of eggs produced per female mosquito per day, φ), the pupa to larva transition rate (ω_p_), the per-bite probability of infection from mosquito to human (pMI), the extrinsic incubation rate (the time required between a mosquito biting an infected host and becoming infectious, ε), and the larva maturation rate (ω_l_) (Table 2). The relationship between these parameters and temperature is well captured by a Briere function [30, 31], which has the form:

**Table 2:** Summary of climate-dependent model parameters related to dengue transmission in mosquitoes. T, W, and H within parentheses indicate that the parameter is a function of temperature, rainfall, and humidity. The traits fit to a Briere function have T_0_ as the critical thermal minimum, T_m_ as the critical thermal maximum, *c* as the scale parameter, and *d* as the shape parameter.

| Symbol | Meaning (code variable) | Value or expression | Source |
| --- | --- | --- | --- |
| $\Phi$ | Intrinsic oviposition rate per female mosquito (EFD) | Briere function dependent on $T$:<br>$\phi(T) = c T (T - T_0)(T_m - T)^{1/d}$ | Function parameters fitted to data from [31] |
| q | Fraction of eggs that hatch to larvae (FEL) | $q(W) = \frac{q_1 W}{q_0 + q_1 W} + q_2$ | [10, 33] |
| Z | Density-dependent reduction factor for egg hatching | $Z = 1 - \frac{N_l}{K}$<br>Where $N_l$ is the total female larval population at a given time and $K$ is total larval carrying capacity of available breeding sites | [10] |
| K | Total larval carrying capacity of breeding sites | $K = N_h * kf * k$<br>Where $N_h$ is the total human population at a given time | [49] |
| kf | Carrying capacity factor (kf) | 6 | Calibrated |
| k | Carrying capacity for larvae per breeding site | $k(W) = \frac{k_0 W}{k_1 + W} + k_2$<br>Where $k_0 = 5$, is the capacity of rain to produce new breeding sites, $k_1 = 30$, is the critical amount of rain required to form breeding sites, $k_2 = 0.1$, is the rain independent variation in the number of breeding sites | [49] |
| a | Biting rate | Briere function dependent on $T$:<br>$a(T) = c T (T - T_0)(T_m - T)^{1/d}$ | Function parameters fitted to data from [31] |
| $\omega_l$ | Per-capita transition rate from larva to Pupa (LMR) | Briere function dependent on $T$: | Function parameters fitted to data from [49] |
| | | $\omega_l(T) = c T (T - T_0)(T_m - T)^{1/d}$ | |
| $\omega_p$ | Per-capita transition rate from pupa to adult (PMR) | Briere function dependent on $T$:<br>$\omega_p(T) = c T (T - T_0)(T_m - T)^{1/d}$ | Function parameters fitted to data from [49] |
| $\epsilon$ | Extrinsic Incubation Rate (PDR) | Briere function dependent on $T$:<br>$\epsilon(T) = c T (T - T_0)(T_m - T)^{1/d}$ | Function parameters fitted to data from [11] |
| pMI | Probability of mosquito infection per bite on an infectious host (pMI) | Briere function dependent on $T$:<br>$pMI(T) = c T (T - T_0)(T_m - T)^{1/d}$ | Function parameters fitted to data from [31] |
| pIM | Probability of human infection per bite by an infectious mosquito;<br>Function of temperature (pIM) | Briere function dependent on $T$:<br>$pIM(T) = c T (T - T_0)(T_m - T)^{1/d}$ | Function parameters fitted to data from [31] |
| $\mu_v$ | Per-capita mortality rate of female adults (muv) | $\mu_v(T, H) = \frac{1}{c(T-T_0)(T-T_m)} + (1 - (0.01 + 2.01 * H)) * g \quad H < 1$<br>$\mu_v(T, H) = \frac{1}{c(T-T_0)(T-T_m)} + (1 - (1.22 + 0.27 * H)) * g \quad H \geq 1$ | Temperature dependent function parameters fitted to data from [11] |
| $\mu_l$ | Per-capita mortality rate of larvae (mul) | $\mu_l(T, W) = \mu_{lt}(T)\{1 + \mu_{al}(W - W_c)\theta[W - W_c]\}$ | Temperature dependent function parameters fitted to data from [49] |
| $\mu_p$ | Per-capita mortality rate of pupae (mup) | $\mu_p(T, W) = \mu_{pt}(T)\{1 + \mu_{ap}(W - W_c)\theta[W - W_c]\}$ | Temperature dependent function parameters fitted to data from [49] |

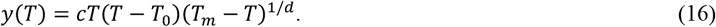

Other parameters depend on multiple climate variables. The adult *Ae. aegypti* mosquito mortality rate (µ_v_) is assumed to be a function of temperature (T) and humidity (H) [11]. The larva (µ_l_) and pupa mortality rates (µ_p_) are assumed to be functions of temperature and rainfall (W) [32]. Finally, the fraction of eggs that hatch to larva (q) and the density-dependent reduction factor (regulating larva population due to competition of limited resources, Z) are assumed to be functions of rainfall only [10, 33]. The following equations describe the functional relationships between the parameters and climate variables:

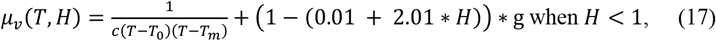

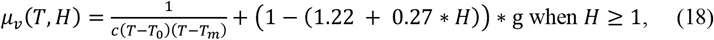

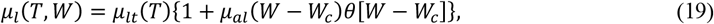

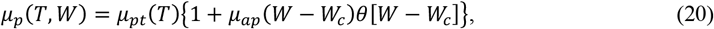

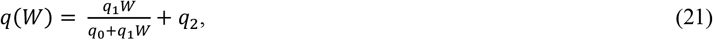

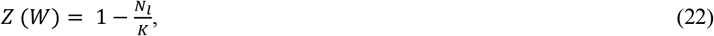

where, *KK* = *NN*_*h*_ * *kkkk* * *kk* and 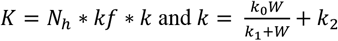

We extracted empirical data pairs for specific mosquito parameter values and their corresponding climate variables from published literature and performed model fitting to estimate the values for function parameters. The data extraction and fitting process, the fitted curves and associated uncertainty are explained in the Supplementary Material (section 1). Setting-specific model parameters requiring calibration and fixed model parameters are described in the following section.

### Model assumptions and calibration

To simulate dengue transmission using the model, we made several (default) assumptions based on the available data, previous publications, and biological plausibility. We used hourly timesteps to capture the rapid dynamics of dengue transmission, the mosquito lifecycle, and the impact of daily diurnal temperature variations on the mosquito population [12]. Since hourly temperature data were not available, we assumed a sinusoidal fluctuation in temperature throughout the day to produce hourly temperature estimates from the available daily temperature data. Instead of daily rainfall we inputted the 14-day average to reflect the pooling of water and maintenance of mosquito breeding sites over many days. We also evaluated alternatives to the default assumptions related to temperature, rainfall, and humidity data processing, which showed that our selections led to more realistic model outputs (Supplementary Material section 2). We assumed a positive net influx of susceptible humans into the population given the net migration of people into Dhaka from other parts of Bangladesh with a lower seroprevalence of dengue (in the order of 1/8^th^ the prevalence in Dhaka [22]. Finally, an initial female mosquito to human population ratio of 2 was used, as it has previously been shown that varying this ratio between 1 and 3 results in similar dynamics of dengue [11, 15].

Due to substantial under-reporting of dengue cases for Dhaka we were unable to calibrate the model directly to time-series data. Instead, we calibrated the model to the seroprevalence estimates for 2014 in Dhaka, the seasonality, and yearly outbreak patterns of the case data (we assumed that these were robust to the under-reporting and reflected the actual seasonality and yearly outbreak patterns). To do this we manually varied three key input parameters that have the largest effect on transmission: the carrying capacity factor (kf), and the two probabilities of transmission from mosquitoes to humans and from humans to mosquitoes using multiplicative factors (β_im_ and β_mi_). The carrying capacity factor, the carrying capacity for larva per breeding site (k), and the total human population (N_h_) available at a given time determined the maximum number of larva that the environment can sustainably support, also known as the total environmental larval carrying capacity (K) (Table 2).

### Model simulations and analysis

Base model simulations were performed using the median values of the model parameters (Tables 1, 2, and 3), the best fit function parameters (Tables S1 and S2), and the default assumptions. We also separately assessed the effect of our default assumptions for individual climate and demographic variables by simulating the best fit parameters but with an alternative assumption. Comparative results for the need for two dengue serotypes and the impact of migration into Dhaka on dengue transmission are presented here, while the comparative results of other assumptions are presented in the Supplementary Material (section 2).

**Table 3:** Setting specific parameters inputted to initiate and then run the dengue transmission model for the period 1995-2014.

| Setting specific parameter | value | Source |
| --- | --- | --- |
| Initial human population size ($N_h$) | 8,332,000 | [24] |
| Immigration rate ($\Omega_{im}$) | 0.0336% of human population/day | [23] |
| Emigration rate ($\Omega_{em}$) | 0.02856% of human population/day | Calibrated to match reported net migration rate [23] |
| Existing seroprevalence in 1995 (ES) | 0.54% | [50] |
| Seroprevalence among incoming individuals (SF) | 1/8<sup>th</sup> of population seroprevalence at any time point | Seroprevalence outside Dhaka was reported to be about 1/8<sup>th</sup> of that in Dhaka in 2014 [22] |

To ascertain the uncertainty surrounding the model’s output, we ran an additional 999 simulations of dengue transmission in Dhaka for the period 1995-2014, with each simulation utilizing one of the 999 sets of parameter values. For the climate-independent parameters, we randomly sampled 999 values from their respective distributions to prepare 999 parameter sets (Table 1). For the climate-dependent mosquito parameters (Table 2), we randomly sampled corresponding values of each function parameter from their respective ranges (Supplementary Material section 1). For the remaining location-specific parameters listed in Table 3, we used fixed values. The output from the 1,000 simulations, including the base model simulation, were then summarized and presented as median and interquartile range (IQR).

## Results

With the default assumptions for individual climate and demographic variables, the model produced repeating annual seasonal outbreaks of dengue between 1995 and 2014 (Figure 3). For the 1995–2014 period, the median annual number of new dengue infections was 1.6 million (IQR: 0.6–2.3 million) and the median of the annual daily peak size (maximum number of new daily infections in a year for the 20-year period) of dengue infections was 24,121 (IQR: 7,807-39,308) across the 1,000 simulations. The median total number of new dengue infections over the 20-year period was 36.5 million (IQR: 33.2–39.1 million). These model results align with estimates from another study [22]. Dengue infections occurred in the model during August to December with the median peak of dengue infection occurring in October. However, the highest number of reported dengue cases in Dhaka peaked in September (Figure 1B), a month before the modelled peak, which tends to be more pronounced (Figure 3E). The model estimated a median seroprevalence in 2014 for Dhaka of 62% (IQR: 58–75%) which is well within the recorded range [22].

**Figure 3:**
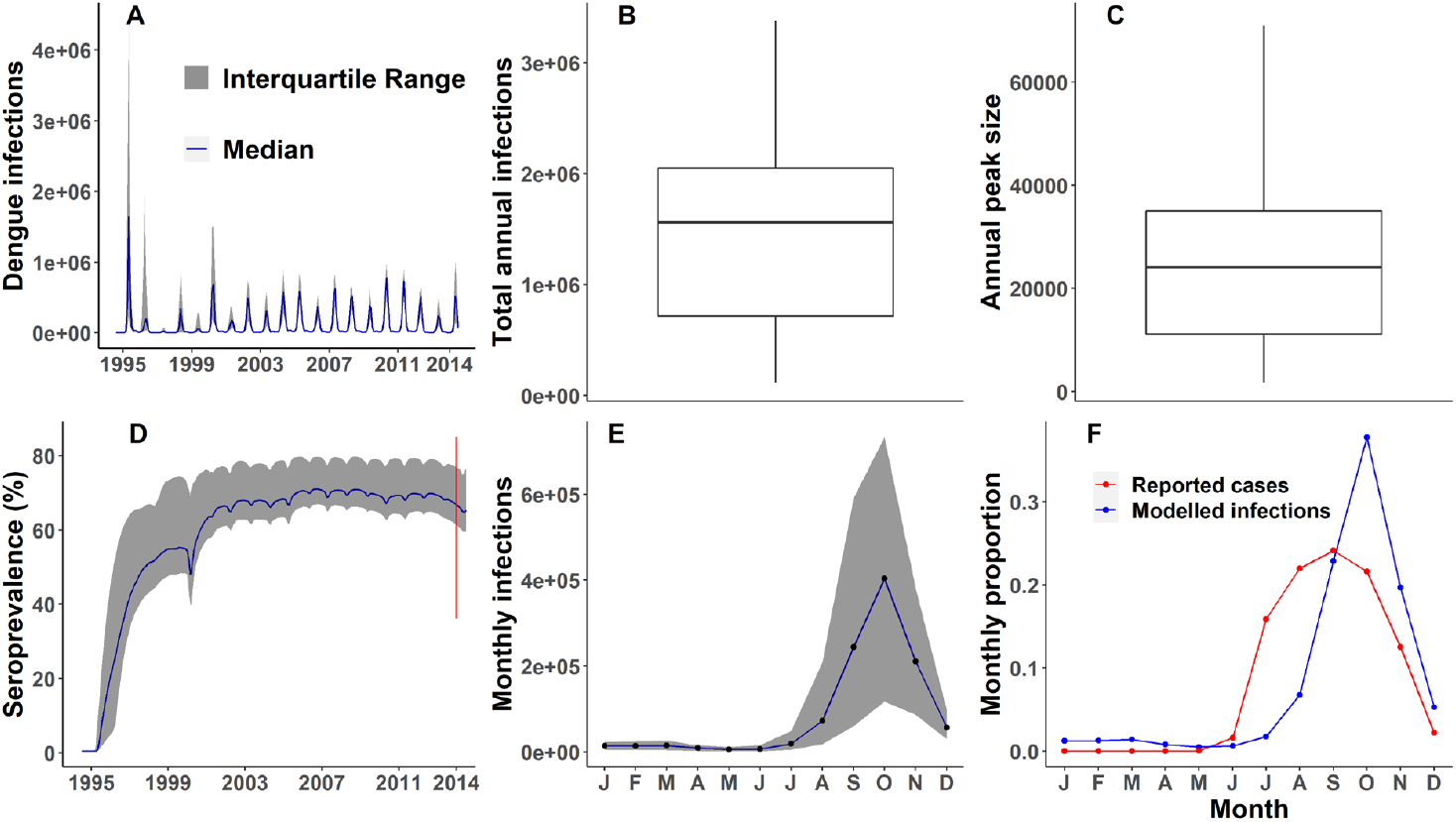
Modelled dengue transmission in Dhaka, Bangladesh (1995-2014): output from 1,000 simulations - (A) Median number of infections and IQR of monthly dengue infections. (B) Distribution of total annual infections across the period - the lower boundary of the box corresponds to the first quartile, the upper boundary corresponds to the third quartile, the line within the box represents the median, the whisker extended from the edges of the box indicate the range of the data, outliers excluded (1,159 outliers = 5.6% of values, maximum value: 9.9e+6). (C) Distribution of annual peak (maximum number of new daily infections each year) across the period, outliers excluded (1,631 outliers = 8.1% of values, maximum value: 5.6e+5). (D) Median seroprevalence and IQR: seroprevalence is defined as the proportion of all individuals ever exposed to any of the two dengue serotypes; red line segment – estimated range of dengue seroprevalence in Dhaka reported in a community-based seroprevalence study [22] (E) Seasonal distribution of dengue infections showing the median (solid line) and IQR (shaded area). (F) Proportion of median number of reported dengue cases and modelled dengue infections in each month.

### Two vs one dengue serotype

The introduction of a single dengue serotype into a population typically took around one year to generate a significant number of infected individuals with potential to result in a large-scale epidemic in the following year (Figure 4). In this scenario, the transmission of dengue continued at a lower level for approximately six years before the next major epidemic. During this period, the susceptible population gradually increased.

**Figure 4:**
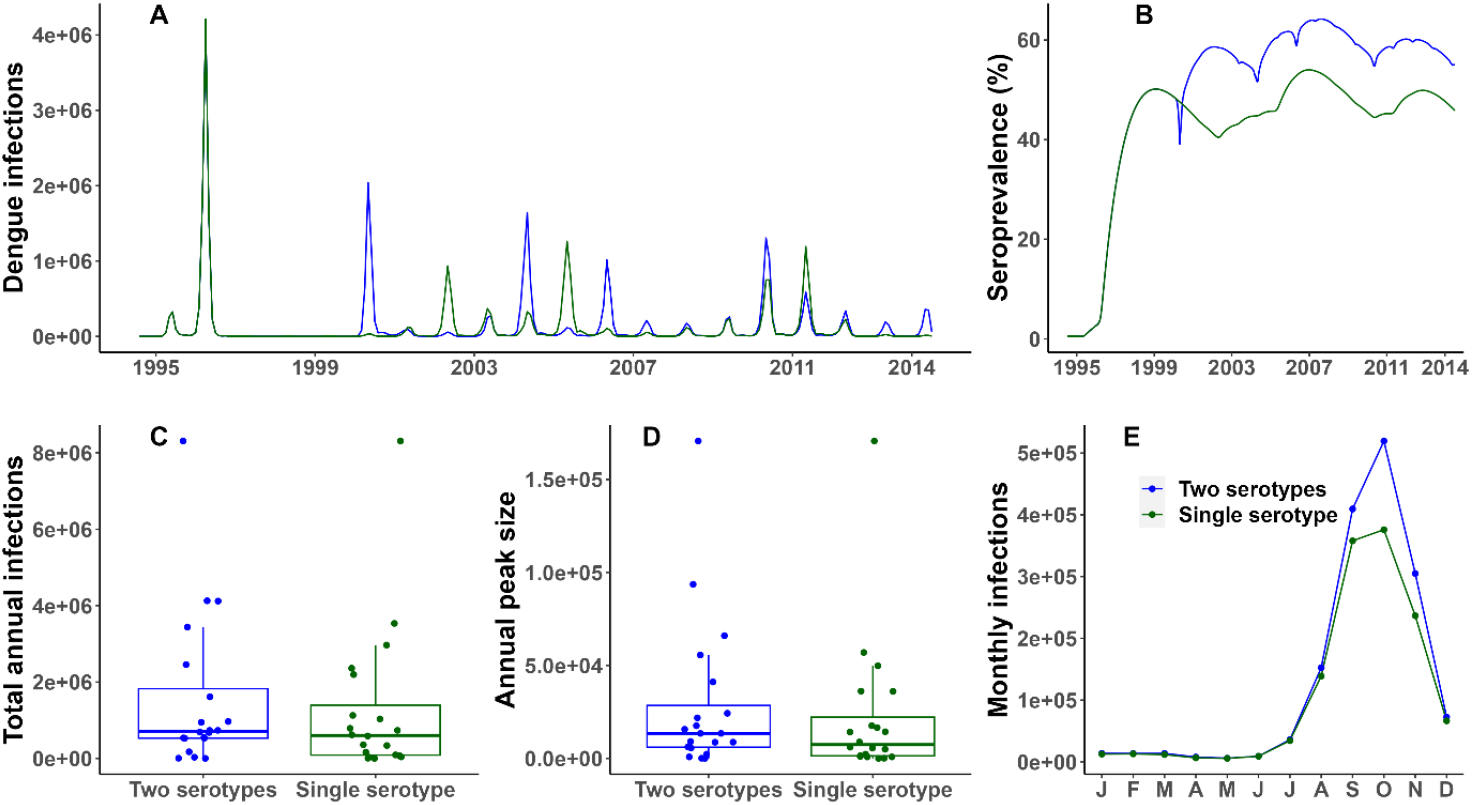
Comparison of model output using two (base-case) vs one dengue serotype (1995-2014) (A) Daily dengue infections. (B) Seroprevalence (%) of dengue. (C) Distribution of annual number of dengue infections: median for a single dengue serotype – 4.19e+5, median for two dengue serotypes – 4.53e+5. (D) Distribution of annual peak number of infections: single serotype median – 6.36e+3, Two serotypes median – 6.52e+3. (E) Seasonality of dengue shown as the mean monthly infections over the years. The single serotype dengue transmission model was not re-calibrated.

The inclusion of a second dengue serotype had a sizable impact on the overall epidemiology of the disease. Following the introduction of the second serotype into the population, five years after the first serotype, another substantial epidemic occurred. However, the size of this epidemic was smaller than the initial outbreak (4.1 vs 8.3 million infections at the peak) that followed the introduction of the first serotype. This decrease in magnitude can be attributed to the presence of cross-immunity in the population, resulting in a reduced number of susceptible individuals. The transmission of two serotypes also led to a higher overall incidence of dengue infections in comparison to the scenario with a single serotype and subsequently produced a higher seroprevalence after twenty years of simulation (55% vs 45% in 2014). Transmission of two dengue serotypes also leads to cyclical changes in the dominant dengue serotype. Furthermore, the double-serotype model resulted in earlier infections during the year with more pronounced monthly mean numbers compared to the single-serotype model, which better aligns with available monthly case data.

### Role of migration

A net inflow of susceptible hosts into the population significantly impacted dengue transmission overall, compared to the no net migration scenario. With net immigration, dengue epidemics started off smaller but escalated later, while zero net migration resulted in an opposite trend, with larger epidemics initially followed by smaller ones (Figure 5). The yearly number of dengue infections (average over 20 years 1,290,819 vs 521,493) and the yearly peak in daily infections also become much higher (average over 20 years 23,027 vs 9,940) compared to when there was no net migration. In the absence of immigration, dengue seroprevalence rapidly reached its peak and stayed high (79% vs 43% in 2014) due to a reduced number of susceptible individuals entering the population, thus reducing overall transmission.

**Figure 5:**
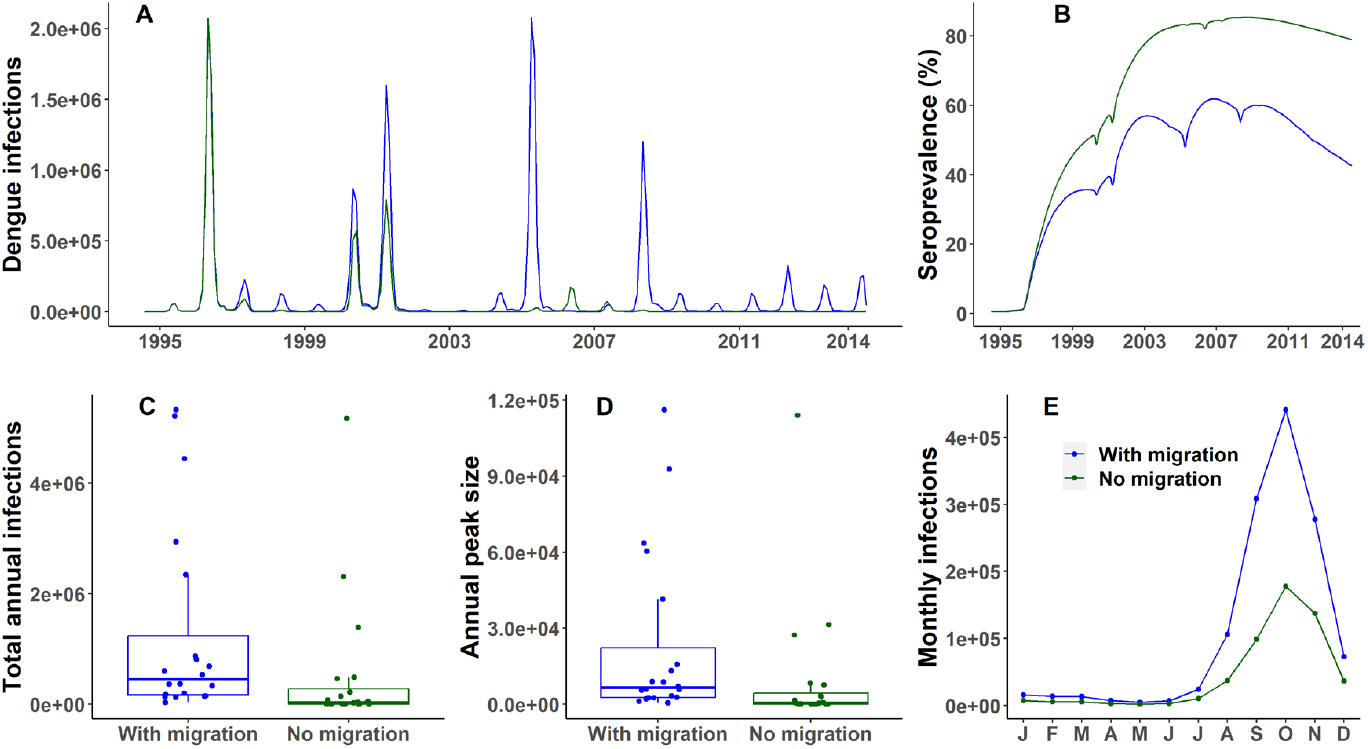
Comparison of model output with net inflow of susceptible hosts (base case) vs no net migration (1995-2014) (A) Daily dengue infections. (B) Seroprevalence (%) of dengue (C) Distribution of annual number of dengue infections: median with migration – 4.53e+5, median without any migration – 3.38e+4. (D) Distribution of annual peak number of infections: median with migration – 6.52e+3, median without any migration – 4.95e+2. (E) Seasonality of dengue shown as the mean monthly infections over the years.

## Discussion

We developed a dengue virus transmission model incorporating the key characteristics of mosquito vectors, the human host population, and their interactions with the dengue virus. The aim was to develop a model suitable for assessing the impact of future climate change on dengue transmission. The model explicitly represents both aquatic and adult phases of the mosquito vector lifecycle, which have different sensitivities to climate. Importantly, the model incorporates two different dengue serotypes and cross-immunity, allowing us to assess the impact of climate changes over the long term. We incorporated the impact of all three climate variables thought to be important for mosquito biology and behaviour in our model (i.e., temperature, rainfall and humidity). We applied this tool to Dhaka, Bangladesh between 1995-2014 when dengue case data and meteorological data were available.

Our model produced recurring yearly dengue outbreaks over a 20-year period when applied to the population in Dhaka, consistent with the repetitive pattern observed in annual dengue case reports. The median annual reported number of dengue cases for the period 2000-2014 is substantially lower than the number of infections estimated by our model [18]. The large discrepancy between reported case numbers and the estimated number of infections is most likely due to a combination of factors: most dengue infections are asymptomatic; not all symptomatic cases seek care in a formal healthcare setting; and only a small proportion of symptomatic dengue cases are confirmed by testing and included in the case-count, with less than 3% of symptomatic dengue cases in Dhaka being reported through the passive surveillance system [19, 34].

In terms of seasonality, the model produced outbreaks aligned with the dengue season in Dhaka but the outbreak peak in the model output occurred a month later during October with the overall simulated outbreak having sharper peak than for the observed data. This disparity could be due to several uncertain factors that were not accounted for but could be considered in future model development. These include the implementation of mosquito vector control measures and awareness campaigns by city health authorities in response to reported dengue cases, resulting in a gradual decrease in case numbers [35] and a potential lag effect of climate variables. Several studies have shown a correlation between temperature and rainfall fluctuations and the dengue peak occurring later [5, 36]. However, the results are inconsistent and the biological mechanism for this lag is unknown.

The model output also showed cyclical changes in the dominant dengue serotype due to the build-up of immunity in the population which is consistent with serotype data for Dhaka, with outbreaks initially driven by DENV 3 (2000-2002) but then predominantly by DENV 1 and DENV 2 in 2013-2016 [37]. DENV 3 re-emerged as the most prevalent serotype in a large outbreak in 2019 and continued to be the major serotype in the following years [38]. The model also showed that the introduction of a new dengue serotype, for which there is little to no population level immunity except for cross-immunity, can lead to more frequent and more pronounced epidemics, as seen in recent years in Dhaka [39].

We found the other important driver of recurring yearly dengue outbreak is immigration, resulting in an increase in the susceptible pool available for dengue infection and further propagation. Caldwell *et al*. arbitrarily assumed immigration of 1% of the study population into the susceptible pool each day [11]. However, this is unrealistic for Dhaka, and we assumed immigration of 0.0336% of the total population per day based on previous data on migration within Bangladesh [23]. Given the significant impact of immigration rate on dengue epidemiology, it is crucial to carefully account for this when modelling dengue for a specific location.

There are limitations to our dengue transmission modelling approach, which must be considered when interpreting our results. First, there is limited data to inform the temperature-mosquito relationships, with no data for Bangladesh, and there could be country specific mosquito traits that have evolved for local climates. Additionally, we did not incorporate the age structure of the host population into our model. However, this is more relevant for models evaluating the potential impact of dengue vaccination, which is more focused on individuals, rather than understanding and comparing the effects of climate change at a population level. Population movement within Dhaka might also be an important driver for disease spread given the heterogeneity in dengue seroprevalence across the city and limited flight range of mosquitoes [40]. The distribution of businesses and industries across Dhaka is such that workers move around the city, leading to a well-mixed population [41]. As a result, the application of our dengue transmission model, which covers the entire city of Dhaka, is appropriate for this context and provides a good overall picture of dengue at the population level. Finally, changes in health policy or interventions targeted at dengue control and prevention in Dhaka were not considered in our study. However, the model is flexible enough to incorporate these factors. As outlined in the methods section, limited epidemiological data on dengue makes it difficult to ensure the robustness of the model estimates. In situations where extensive laboratory and hospital-based surveillance is logistically challenging, periodic seroprevalence surveys could provide valuable data for developing robust estimates with the model. Despite the limitations, our model has a good balance between complexity and simplicity, providing insight of dengue dynamics at the population level.

In this paper, we developed a climate-driven dengue virus transmission model, which incorporates the mosquito vector life cycle and temperature, rainfall, and humidity variables simultaneously and factors that shape long-term dengue epidemiology, including cross-immunity to different dengue serotypes and demographic shifts resulting from population migration. We successfully applied this model to Dhaka, accurately reflecting the available dengue data through the variation in climate variables. The model is applicable for other settings where climate data and dengue epidemiological data are available. By accounting for climate variables, host immunity, and their interplay, our model is well-suited for evaluating the long-term impact of climate change under different scenarios.

## Data Availability

All data and model code are available online at https://github.com/kkpaul-ide/Dengue_Climate_Model

