## Supplementary Material for "A Climate-Driven Mechanistic Transmission Model to Characterize Dengue Epidemiology in Dhaka, Bangladesh"

This supplementary material provides further details on the empirical data extraction from published literature, the fitting of mosquito parameters to this data, and climate data processing. We also provide additional comparative results for the assumptions related to climate variables and the female mosquito to human population ratio. All parameter fitting and model code are available under an open access license at the following URL : https://github.com/kkpaul-ide/Dengue_Climate_Model [1].

#### 1. Climate-dependent model input parameters

##### 1.1 Parameters dependent on temperature only

Experiments have established that several model parameters related to the behaviour and lifecycle of mosquitoes are affected by temperature: the biting rate (a), the per-bite probability of infection from human to mosquito (pIM), the oviposition rate (φ), the pupa to larva transition rate ($\omega_p$), the per-bite probability of infection from mosquito to human (pMI), the extrinsic incubation rate (ε), and the larva maturation rate ($\omega_l$) [2]. The oviposition rate is the rate at which female mosquito vectors lay their eggs, also described as eggs per female mosquito per day in previous modelling studies [3]. Previously, Yang *et al.* utilized polynomial fits to the data obtained from laboratory experiments to analyse the relationships between these parameters and temperature [2]. While polynomials provide good fits to data, higher degree polynomials are unlikely to describe the causal relationships between temperature and mosquito parameters because they do not capture the underlying biology mechanistically and are prone to overfitting. Another study by Mordecai *et al.* fitted the thermal responses of biting rate, infection and transmission probabilities, oviposition rate, and extrinsic incubation rate using a Briere function and a Bayesian fitting procedure but did not report the final fitted coefficients [4]. We therefore choose to re-fit Briere functions to the relationships using the published data. Briere functions reflect an asymmetric unimodal thermal response. Such functions are of the following form (equation 1) and provide a simple but biologically plausible nonlinear description of temperature-dependent mosquito parameters [5]

$$y(T) = cT(T - T_0)(T_m - T)^{1/d}. \quad (1)$$

The critical thermal minimum and maximum are represented by $T_0$ and $T_m$, respectively, while $c$ is the scale parameter and $d$ is the shape parameter.

We used WebPlotDigitizer, a web-based tool, to extract empirical data pairs of the relevant mosquito parameter and temperature values from published books and papers [2, 6, 7], to perform our own fitting of Briere functions and obtain parameter estimates for the model. The number of data pairs for the seven mosquito parameters, ranged from 7 to 23 and these are the same data sources as used by Mordecai *et al.* [4].

To fit a Briere function to the extracted data for each parameter, we used non-linear least squares (NLS) with a bootstrapping technique to produce 95% confidence intervals (CI). As some of the extracted datasets did not cover the full temperature range for mosquito survivability, we imposed constraints using assumed values for the minimum and maximum temperature in the Briere function to ensure the fitting produced realistic curves based on the visual inspection of the Mordecai *et al.* fits and data from other empirical studies [4, 8]. Using the 95% CI for the underlying parameters from the initial NLS fit to sample and produce the uncertainty range for the mosquito parameter produces unrealistically large ranges in the estimated values. We therefore employed a bootstrapping technique to produce uncertainty ranges for the mosquito parameters used in the model. This involved creating multiple resampled datasets from the extracted original dataset and then randomly selecting a new data set from the original data with replacement and then performing the NLS fitting. One issue with this method is that the NLS fitting algorithm used may not converge for all bootstrapped data sets. To account for this, we resampled the extracted original dataset 25,000 times with replacement, fitted the Briere function to each parameter set, and then randomly selected 1,000 parameter sets from the sets where a fit was achieved. From the 1,000 sets of parameters obtained we were able to obtain a robust estimate of the uncertainty of the temperature dependent parameters for input into the dengue model. To conduct the non-linear least squares fitting we used the R package “minpack.lm” with the “tidymodels” package to do the bootstrapping [9, 10]. Fits produced using this method are shown in Figure S1 and the associated code is accessible at the project repository [1].

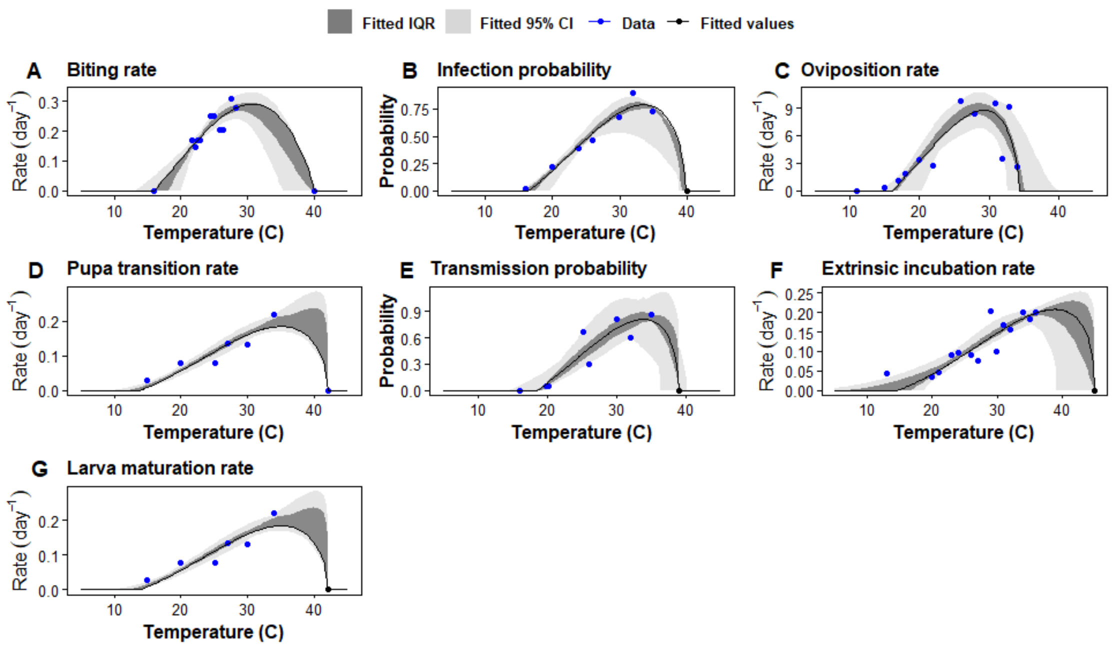


**Figure S1: Briere function fits of the thermal responses of *Ae. aegypti* behaviour and lifecycle traits**: (A) Biting rate ($a$); (B) Infection probability ($pIM$); (C) Oviposition rate ($\phi$); (D) Pupa transition rate ($\omega_p$); (E) Transmission probability ($pMI$); (F) Extrinsic incubation rate ($\epsilon$); (G) Larva maturation rate ($\omega_l$). The black line represents the best fit, the darker and lighter shaded bands around the fitted values denotes the interquartile range (IQR) and 95% Confidence Interval (CI) from the 1,000 fitted curves from the bootstrapping process. Blue dots are the extracted data points from Yang *et al.* [2] and black dots are the assumed data points used to constrain the curve to remain in a biologically plausible range, based on visual inspection of the fit by Mordecai *et al.* [4].

#### 1.2 Parameters dependent on temperature and humidity

Temperature directly influences adult *Ae. aegypti* mosquito mortality rate ($\mu_v$) and humidity play an indirect role, with its impact depending on the temperature [11, 12]. Caldwell *et al* fitted a quadratic function to the available empirical data on thermal component of adult mosquito mortality but did not report the fitted coefficients [3]. A quadratic function provides a simple and realistic functional form for describing these parameters. Hence, as described above, we used WebPlotDigitizer to extract empirical data pairs and performed fitting for the thermal component of the mortality function. Using the same fitting procedure described above, we obtained a robust estimate for the value and the uncertainty of the function parameters. The number of data points and the nature of the relationship meant we did not need to implement constraints for the fitting process (see Figure S2 C).

The impact of humidity is incorporated into the adult mosquito mortality rate function linearly, as described by Caldwell *et al.* [3] using the following equations:

$$\mu_v(T,H) = \frac{1}{c(T-T_0)(T-T_m)} + \big(1 - (0.01 + 2.01 * H)\big) * y \quad H < 1, \qquad (2)$$

$$\mu_v(T,H) = \frac{1}{c(T-T_0)(T-T_m)} + \big(1 - (1.22 + 0.27 * H)\big) * y \quad H \geq 1, \qquad (3)$$

where the rate constant ($c$), minimum temperature ($T_0$), and maximum temperature ($T_m$) are estimated from the fitting process (Table S2), and $y$ is a scaling factor that is set to 0.005 and

0.01, respectively, to restrict mosquito mortality rates within the range of mortality rates estimated by other studies [4, 11].

##### 1.3 Parameters dependent on temperature and rainfall

Empirical studies have shown larva mortality rate ($\mu_l$) and pupa mortality rate ($\mu_p$) are dependent on temperature. Rainfall also affects larva and pupa mortality because during heavy rainfall, mosquito breeding sites can be emptied of larva and pupa through flushing, a process known as physically induced mortality. For the thermal component of larva and pupa mortality, we chose to fit a quadratic function to the published experimental data. Yang *et al.* fitted empirical data on the relationship between temperature and mortality rates of larva and pupa using high order polynomials which are biologically implausible [13]. As for adult mosquito mortality rate, we extracted experimental data and conducted quadratic fitting. We then employed a bootstrapping technique to quantify the uncertainty of the parameters in the model. Physically induced mortality due to flushing is included as additional mortality in the larva (equation 18) and pupa (equation 19) mortality equations:

$$\mu_l(T, W) = \mu_{lt}(T)\{1 + \mu_{al}(W - W_c)\theta[W - W_c]\}c, \tag{4}$$

$$\mu_p(T, W) = \mu_{pt}(T)\{1 + \mu_{ap}(W - W_c)\theta[W - W_c]\}. \tag{5}$$

Here, $\mu_{lt}$ and $\mu_{pt}$ are temperature-dependent mortality rates fitted to experimental data (Figures S2A and S2B). The function parameters $\mu_{al}$ and $\mu_{ap}$ are additional mortality for larva and pupa due to heavy rain and are set to be 0.001 per mm rainfall as per Liu-Helmersson *et al.* [14]. $W_c$ is the critical rain volume required for flushing of mosquito breeding sites. $\theta(x)$ is the Heaviside function [15], that is, $\theta(x) = 1$, if $x \geq 0$ and is otherwise $\theta(x) = 0$.

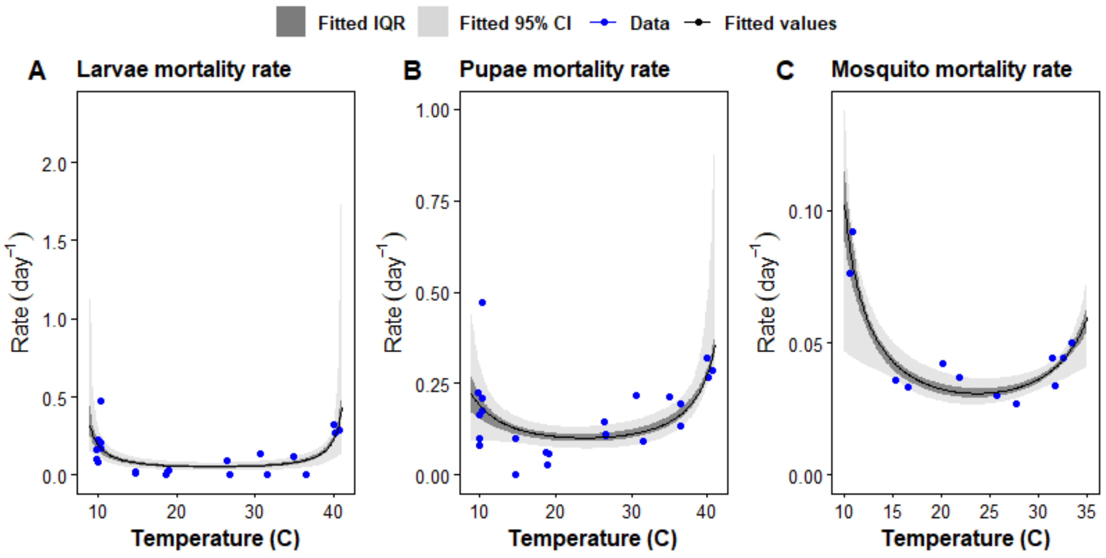


**Figure S2: Quadratic fit of the thermal responses of mortality rates of three lifecycle stages of *Ae. aegypti***: (A) Larvae mortality rate ($\mu_{lt}$); (B) Pupae mortality rate ($\mu_{pt}$); (C) Mosquito mortality rate ($\mu_v$). The darker shaded band around fitted values denotes the IQR from the 1000 fitted curves from the bootstrapping process. The lighter shaded band around fitted values denotes the 95% CI from the 1,000 fitted curves obtained in the bootstrapping process. Data points here represent empirical observations of mortality in the absence of rain.

#### 1.4 Parameters dependent on rainfall only

Larval population is regulated by the fraction of eggs that hatch to larva ($q$) and the density-dependent reduction factor ($Z$). These parameters are dependent on rainfall [14, 15]. The rain-dependent function for the fraction of eggs that hatch to larva ($q$) is given by:

$$q(W) = \frac{q_1 W}{q_0 + q_1 W} + q_2, \quad (6)$$

where W is the amount of rain in millimetres (mm); $q_0$ = 0.2 mm/day is the critical amount of rainfall to promote hatching (the lower $q_0$, the higher the influence of rainfall on hatching); $q_1$= 0.02 is the capacity of eggs hatching with rainfall, 2% egg hatching with every mm of rainfall; and $q_2$ = 0.037 is the hatching fraction from rain-independent breeding sites, e.g., household containers used for water storage.

As in Liu-Helmersson *et al.* [14], the density-dependent reduction factor ($Z$) for egg hatching is given by:

$$Z = 1 - \frac{N_l}{K}, \quad (7)$$

where $N_l$ is total female larval population at a given time and $K$ is total larval carrying capacity of all available breeding sites. The parameter Z ranges between 0 and 1. When Z approaches 1, it indicates that the total larval population is close to the maximum carrying capacity. Conversely, when the total larval population increases while the total larval carrying capacity remains static, Z approaches 0, suggesting that the population is far from reaching its maximum capacity.

The total larval carrying capacity of available breeding sites is then given by:

$$K = N_h * kf * k, \quad (8)$$

where $N_h$ is the total human population at a given time; $kf$ is the Carrying capacity factor; and $k$ is the carrying capacity for larvae per breeding site. The function parameter $k$ is dependent on rainfall as given by:

$$k(W) = \frac{k_0 W}{k_1 + W} + k_2, \quad (9)$$

where $k_0$ = 5 is the capacity of rainfall to produce new breeding sites, that is 5 new breeding sites established per mm of rainfall; $k_1$ = 30 mm/day is the amount of rainfall to produce half of the maximum breeding sites (the lower $k_1$, the higher the influence of rainfall on increasing breeding sites); and $k_2$ = 0.1, is the contribution from rain independent breeding sites.

#### 1.5 Climate data processing

When sub-daily temperature observations are not available, model simulation studies of dengue often use the mean daily temperature for each time-point during a day as a simplifying assumption [3, 16]. Using the mean daily temperature as a proxy assumes that mosquitoes are active continuously throughout the day and night, allowing for transmission to occur at any time rather than during specific periods. To overcome this limitation, we processed the climate variables to produce estimates for the temperature during the day before calculating climate-dependent transmission parameters. We used the daily mean temperature ($T_{mean}$) and the difference between minimum and maximum temperatures (diurnal temperature range or $DTR$) to generate sub-daily temperature data assuming a sinusoidal variation in temperature over the day ($T_{sinusoid}$) [17, 18]. This is given by the following equation:

$$T_{sinusoid} = T_{mean} + DTR * \frac{\sin\left(0{:}(t_{steps}-1)*pi*\frac{1}{t_{steps}}*2\right)}{2}, \quad (10)$$

where $t_{steps}$ is the number of model time steps per day, 24 in our model. We then used the processed hourly temperature data to calculate transmission parameters dependent on temperature.

To calculate the rainfall-dependent parameters we used average rainfall over the previous 14 days instead of daily recorded rainfall. The average rainfall over the previous 14 days better represents the accumulation of rainwater in outdoor containers required for mosquito egg-laying and propagation.

To calculate the humidity-dependent adult mosquito mortality rate (equation 2 and 3), we used saturation vapour pressure deficit (SVPD) instead of the available observed daily relative humidity values. SVPD is used as it is a more informative measure of the effect of humidity on mosquito survival than relative humidity [11]. SVPD is calculated using the daily relative humidity (RH) and daily mean temperature ($T_{mean}$) using the following equation:

$$SVPD = \left(1 - \left(\frac{RH}{100}\right)\right) * SVP, \quad (11)$$

where $SVP = 610.7 * 10^{7.5*T_{mean}/(237.3+T_{mean})}$. Mosquito mortality as function of SVPD at 25°C temperature is shown in Figure S3, the units of SVPD used in the x-axis of the figure are in kilo Pascals (KPa).

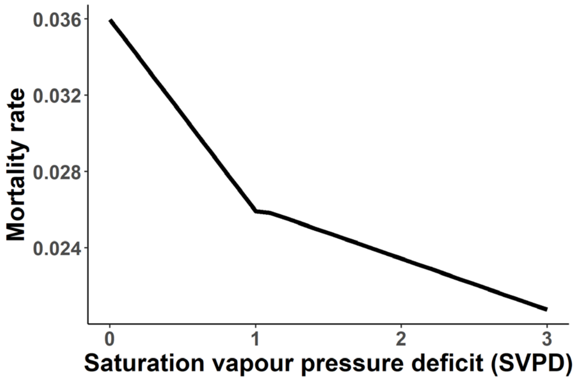


**Figure S3: Adult mosquito mortality rate as a function of saturation vapour pressure deficit (SVPD) when temperature fixed at 25°C**. This linear relationship is a step function where the slope of the relationship is steeper for SVPD $\leq 1$ compared with SVPD $> 1$.

**Table S1:** Fitted Briere function parameters for temperature-dependent *Ae. aegypti* mosquito behaviour and lifecycle traits, best fit values along with IQR in parentheses. The traits fit to a Briere function have $T_0$ as the critical thermal minimum, $T_m$ as the critical thermal maximum, $c$ as the scale parameter, and $d$ as the shape parameter. The units for $T_0$ and $T_m$ are degree Celsius while $c$ and $d$ do not have any units.

| Trait | $\boldsymbol{T_0}$ | $\boldsymbol{T_m}$ | $\boldsymbol{c}$ | $\boldsymbol{d}$ |
|---|---|---|---|---|
| Biting rate ($a$) | 15.94 (15.93, 18.11) | 40.07 (40.07, 40.09) | 8e-5 (4.3e-5, 8e-5) | 1.07 (0.78, 1.05) |
| Infection probability ($pIM$) | 16.45 (15.87, 17.43) | 39.78 (39.24, 39.89) | 5e-4 (5e-4, 5e-4) | 1.81 (1.53, 1.83) |
| Oviposition rate ($\phi$) | 16.06 (15.96, 16.85) | 34.36 (34.32, 35.14) | 9e-3 (5.8e-3, 9e-3) | 1.76 (1.19, 1.71) |
| Pupa transition rate ($\omega_p$) | 13.06 (12.12, 13.51) | 41.75 (41.07, 42.12) | 1e-4 (1e-4, 1e-4) | 0.99 (0.96, 1.03) |
| Transmission probability ($pMI$) | 18.94 (18.43, 19.29) | 39 (39, 39) | 7.9e-4 (5.8e-4, 8e-4) | 2.31 (1.64, 4.42) |
| Extrinsic incubation rate ($\epsilon$) | 14.77 (7.51, 16.73) | 45 (45, 45) | 1.4e-4 (8.9e-5, 1.4e-4) | 2.89 (1.86, 6.07) |
| Larva maturation rate ($\omega_l$) | 13.51 (12.48, 13.91) | 42 (42, 42) | 9.78e-5 (8.9e-5, 1.8e-4) | 1.99 (1.88, 8.22) |

**Table S2:** Fitted quadratic function parameters for the temperature-dependent component of the mortality rates for different lifecycle stages of *Ae. aegypti* mosquito, best fit values along with IQR in parentheses. The traits fit to a quadratic function have $T_0$ as the critical thermal minimum, $T_m$ as the critical thermal maximum, and $c$ as the scale parameter. The units for $T_0$ and $T_m$ are degree Celsius while $c$ has not units.

| Trait | $\boldsymbol{T_0}$ | $\boldsymbol{T_m}$ | $\boldsymbol{c}$ |
|---|---|---|---|
| Larva mortality rate ($\mu_l$) | 7.51 (7.02, 8.07) | 42.10 (41.71, 42.4) | -0.06 (-0.08, -0.05) |
| Pupa mortality rate ($\mu_p$) | 3.62 (0.92, 5.12) | 44.16 (43.46, 44.97) | -0.02 (-0.03, -0.02) |
| Mosquito mortality rate ($\mu_v$) | 7.31 (6.62, 7.81) | 39.99 (39.03, 41.40) | -0.12 (-0.14, -0.10) |

### 2 Additional results

We assessed the effect of our default assumptions for individual climate and demographic variables by simulating the best fit parameters but with an alternative assumption. The results from this simulation were then compared to the best fitting results for the observed period with the default assumption in place to assess the impact of this assumption. Comparative results for the need for two dengue serotypes and the impact of migration into Dhaka on dengue transmission are presented in main text, while the comparative results of other assumptions are presented here. Summary results for different assumptions compared to the base-case are presented in Table S3.

#### 2.1 Impact of climate variables

##### 2.1.1 Mean daily temperature vs sinusoidal temperature change over each day

When simulating the model using the mean daily temperature for each hour of the day compared to a sinusoidal temperature variation between the minimum and maximum temperature each day, we found repeating annual dengue epidemics of different magnitudes (see Figure S4). With a sinusoidal temperature variation, large-scale epidemics were less frequent compared to when mean daily temperature was used for all timesteps in a day. The seroprevalence at the end of 20-year simulation was 60% and 43% for the mean and sinusoidal daily temperature scenarios, respectively, although there was a steeper drop for the sinusoidal temperature scenario in the last five years exaggerating the difference (see Figure S4B). Additionally, the monthly infections tend to rise earlier in the year when the mean temperature is used instead of the sinusoidal temperature. The reason for this difference is due to the sinusoidal approach having periods of the day where the temperature is cooler and further away from the mosquitoes' optimal temperature. However, the sinusoidal approach better represents temperature fluctuations throughout the day and has no impact on simulation time or the complexity of the analyses. Therefore, until further empirical data becomes available, we chose to retain this sinusoidal temperature assumption for our analysis.

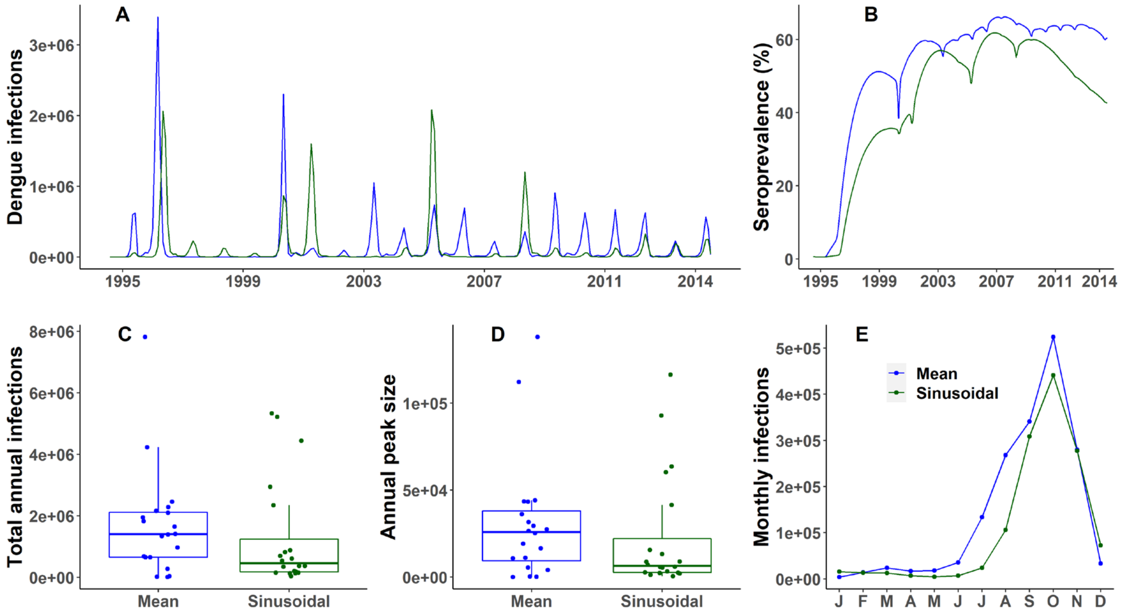


**Figure S4: Comparison of model output using mean temperature vs sinusoidal temperature throughout the day (1995-2014)** - (A) Daily dengue infections. (B)

Seroprevalence (%) of dengue. (C) Distribution of annual number of dengue infections: median for using mean daily temperature – 1.39e+6, median for using sinusoidal distribution of temperature over the day – 4.53e+5. (D) Distribution of annual peak number of infections: median for using mean daily temperature – 2.59e+4, median for using sinusoidal distribution of temperature over the day – 6.52e+3. (E) Seasonality of dengue shown as the mean monthly infections over 1995-2014.

##### 2.1.2 Daily rainfall vs previous 14-day average rainfall

Rainfall impacts multiple mosquito-related parameters in the dengue transmission model. As expected, almost no transmission of dengue happened in our model if there was no rainfall because the parameters describing eggs hatching to larva and the carrying capacity of all breeding containers requires rainfall above a threshold level for mosquito breeding and propagation to occur. Similarly, excessive rainfall attenuated mosquito breeding and propagation reflecting mortality due to aquatic stages of larva and pupa being washed away. Using daily rainfall versus previous 14-day average rainfall as inputs to our model produced similar repeating dengue epidemics over the study period (see Figure S5). The outbreak size in early years was bigger and the total number of dengue infections over the 20-year simulation period was slightly higher if previous 14-day average rainfall is used to run the model. In contrast, the seroprevalence of dengue after 20 years was similar for the two approaches. Model output from both the approaches was consistent with that observed data. This result is similar to what was found by Liu-Helmersson *et al.* [19]. While using the previous 14-day average rainfall in the model to account for accumulated rainwater in containers would intuitively help propagate the mosquito vectors by ensuring breeding sites are maintained over the gestation and pupation periods. Using daily rainfall in the model can be considered a reasonable simplifying assumption for simulation purposes (at least for locations where the variability and seasonality of rainfall are similar to that of Dhaka). However, in settings with different rainfall patterns, such as arid locations, it may be more realistic to utilize the average rainfall over the previous 14 days.

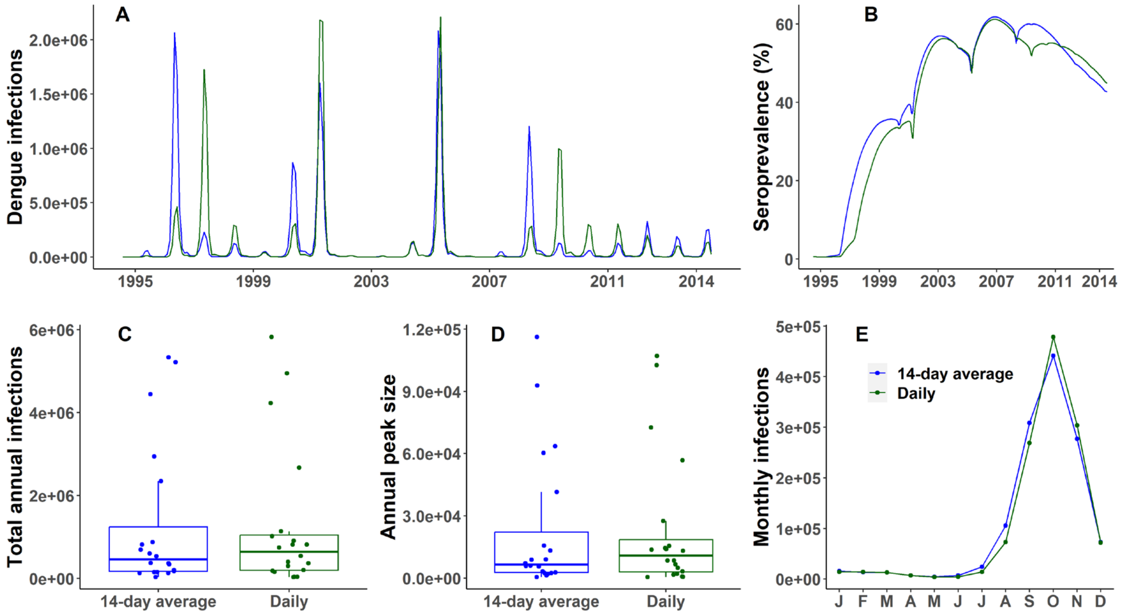


**Figure S5: Comparison of model output using previous 14-day average rainfall vs daily rainfall (1995-2014)** – (A) Daily dengue infections. (B) Seroprevalence (%) of dengue. (C) Distribution of annual number of dengue infections: median for 14-day average rainfall –

4.53e+5, median for daily rainfall – 6.39e+5. (D) Distribution of annual peak number of infections: median for 14-day average rainfall – 6.52e+3, median for daily rainfall – 1.09e+4. (E) Seasonality of dengue shown as the mean monthly infections over the years.

##### 2.1.3 Impact of humidity

Humidity affects the adult mosquito mortality rate parameter in our dengue transmission model. When humidity was excluded from the model, it led to more frequent dengue epidemics that were of a more uniform size compared to when humidity was included. However, incorporating humidity led to explosive epidemics and with longer intervals between them and a slightly higher seroprevalence at the end of 20-year simulation period. It also resulted in a smaller median for the total annual dengue infections and a smaller median size for the annual peak in dengue infections. Additionally, taking humidity into account in the model caused the onset of dengue cases to occur earlier in the year. By incorporating humidity into the transmission model, a better representation of the observed dengue data from Dhaka was achieved, particularly in terms of capturing the explosive nature of dengue epidemics and their seasonality.

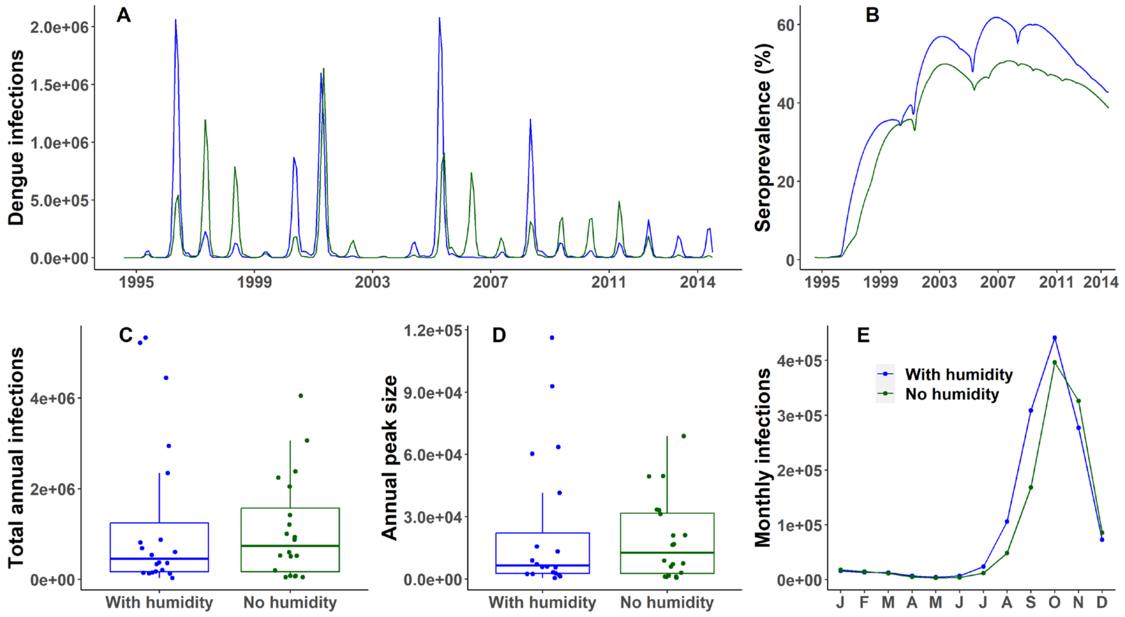


**Figure S6: Comparison of model output with or without incorporation of humidity (1995-2014)** - (A) Daily dengue infections. (B) Seroprevalence (%) of dengue. (C) Distribution of annual number of dengue infections: median when humidity incorporated – 4.53e+5, median when humidity excluded – 7.37e+5. (D) Distribution of annual peak number of infections: median when humidity incorporated – 6.52e+3, median when humidity excluded – 1.27e+4. (E) Seasonality of dengue shown as the mean monthly infections over the years.

#### 2.2 Impact of initial female mosquito to human population ratio

There is a lack of reliable data for the ratio of female mosquito vector to human host population. Our model dynamically monitors both the mosquito and human populations, but we wanted to examine the effect of the initial female mosquito vector to human population ratio on overall dengue transmission. Figure S7 displays a comparison of the model's output when the ratio is set to 4 instead of our assumed 2. It is evident from the figure that the initial ratio had a minimal impact on the overall dynamics of dengue transmission. The ratio of female mosquitoes to the human population fluctuates dynamically in our model due to variations in climate factors, as these factors affect the adult mosquito population in various ways. According to the results of this comparison, the ratio is adaptable to different settings without resulting in significant changes in dengue dynamics.

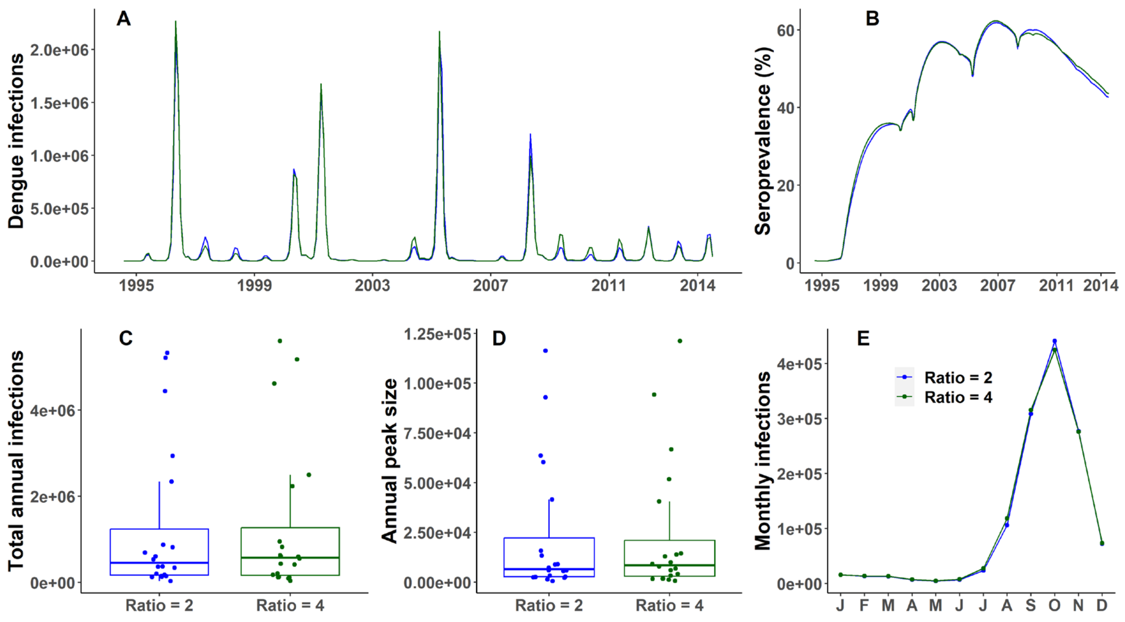


**Figure S7: Comparison of model output with different initial female mosquito to human population ratios (1995-2014)** - (A) Daily dengue infections. (B) Seroprevalence of dengue. (C) Distribution of annual number of dengue infections: median when initial vector population is double of hosts – 4.53e+5, median when initial vector population is four times of hosts – 5.73e+5. (D) Distribution of annual peak number of infections: median when initial vector population is double of hosts – 6.52e+3, median when initial vector population is four times of hosts – 8.44e+3. (E) Mean monthly infections over the period.

**Table S3:** Summary results for different assumptions compared to the base-case, Dhaka, Bangladesh (1995-2014). Median and interquartile ranges (numbers in parenthesis) for annual infections and annual peak (maximum number of new daily infections each year) across the 20-year simulation period shown.

| Scenario | Seroprevalence (%) at the end of 2014 | Annual infections | Annual peak |
|---|---|---|---|
| Base | 43 | 4.53e+5 (1.70e+5, 1.24e+6) | 6.52e+3 (2.69e+3, 2.21e+4) |
| Mean temperature over the day | 60 | 1.39e+6 (6.50e+5, 2.11e+6) | 2.59e+4 (9.46e+3, 3.80e+4) |
| Daily rainfall | 45 | 6.39e+5 (1.96e+5, 1.04e+6) | 1.09e+4 (2.93e+3, 1.85e+4) |
| No humidity | 39 | 7.37e+5 (1.67e+5, 1.57e+6) | 1.27e+4 (2.67e+3,3.18e+4) |
| Initial female mosquito/human ratio = 4 | 44 | 5.73e+5 (1.63e+5, 1.28e+6) | 8.44e+3 (2.90e+3, 2.09e+4) |
